# Glaucoma Detection Using Deep Learning and Prompt-Based Explainable Report Generation

**DOI:** 10.64898/2025.12.13.25342203

**Authors:** Syed Ali Raza Naqvi, Saad B. Ahmed

## Abstract

Glaucoma is a leading cause of irreversible blindness and requires early detection to prevent vision loss. This study proposes a novel framework for automated glaucoma detection using fundus images, integrating deep learning and explainable artificial intelligence (XAI). By unifying five public datasets (RIM-ONE, ACRIMA, DRISHTI-GS, REFUGE, and EyePACS), we have created a diverse dataset to enhance model generalizability. An ensemble of five deep learning models, three convolutional neural networks (ResNet50, EfficientNet-B0, DenseNet121) and two transformer-based models (Vision Transformer, Swin Transformer) are trained for robust classification. Grad-CAM and attention rollout visualizations provided insight into model decision making, highlighting critical regions such as the optic disc and cup. These visualizations, combined with ensemble predictions, were processed by Google Gemini 1.5 Flash to generate clinician-style diagnostic reports. The ensemble model has achieved a test accuracy of 95.38% and an AUC of 0.99, outperforming individual models. This framework improves diagnostic accuracy and interpretability, bridging the gap between AI predictions and clinical utility, with potential for future integration into real-world ophthalmic workflows.

## 1. Introduction

Glaucoma is a prevalent neurodegenerative condition affecting the eye, marked by the death of retinal ganglion cells (RGCs) and the degeneration of their axons [1]. Patients experience a progressive constriction of their visual field associated with an enlargement of the optic disc cupping, often remaining asymptomatic until the condition reaches more advanced stages [2]. Various risk factors are reported including high intraocular pressure (IOP), myopia, aging, thin central corneal thickness (CCT) and corneal hysteresis, family history, oxidative stress, neuroinflammation, low ocular blood flow (OBF), and lifestyle factors such as diabetes, smoking, and diet [3]. In the last 3 decades, there has been a significant increase in the global impact of glaucoma, with the prevalence increasing by 92.53% [4]. Almost 40% of people with absolute glaucoma are unaware of their condition [5].

Artificial Intelligence (AI) is a field of computer science dedicated to the creation of algorithms capable of simulating human intelligence [6]. In the diagnosis of glaucoma, AI serves as a valuable aid in detecting changes in optical coherence tomography scans, visual field tests, and especially fundus images. Fundus images are widely used due to their high availability, good image quality, and cost-effectiveness [7]. Machine Learning (ML) is a branch of AI that allows algorithms to recognize patterns in data, often depending on features that are engineered manually. These involve algorithms such as support vector machines (SVM), decision tree, etc., are used to detect glaucoma but are dependent on the inputs provided by the experts. When there is a need to process large volumes of data, traditional machine learning approaches often fail to provide specialized solutions; this is where Deep Learning (DL) becomes essential. DL learns features directly from raw data through neural networks, especially convolutional neural networks (CNNs), which are particularly efficient in image analysis. Although DL minimizes the need for manual feature extraction, it raises issues related to explainability [8]. The application of AI allows for the quick, accurate and automated detection of glaucoma by examining complex imaging data. This improves early diagnosis in low-resource environments and aids in clinical decision-making [9].

It is very important for healthcare professionals to understand how the AI algorithm made a decision and what steps were taken to reach a decision [10]. To help patients and clinicians better understand how AI makes its decisions, interpretability techniques like Gradient-weighted Class Activation Mapping (Grad-CAM) are increasingly being integrated into medical imaging workflows, offering visual explanations that highlight the most relevant regions influencing the model’s predictions [11]. Explainable AI (XAI) refers to techniques that enhance the transparency and interpretability of AI models, particularly those based on DL, due to their black-box nature. This allows for insights in the decision-making processes of these models, thereby promoting trust and supporting clinical validation [12].

The research presented by numerous researchers in this direction in recent years has made significant progress, but several limitations remain.

- Most existing studies relied on individual datasets, which may lead to overfitting and cause poor generalization by considering varied imaging conditions.
- While interpretability techniques such as Grad-CAM are frequently used to visualize important regions, they are often applied in isolation and not integrated into the final decision-support pipeline, thus, limiting their clinical relevance.
- Vision language models, such as GPT-4V and Gemini, have shown promise in image-based tasks, but they have not been fully explored to generate clinician-style explanations or diagnostic summaries based on model predictions.

This study aims to develop a framework based on deep learning for glaucoma detection, with a strong emphasis on integrating explainability to improve clinical trust and transparency. The study follows a structured approach. A unified dataset is constructed by aggregating data from multiple publicly available sources to ensure diversity and robustness. Explainability techniques, including Grad-CAM and attention-based visualizations, are applied to highlight key anatomical regions that influence model predictions. The model output comprising predicted class, confidence score, and both original and explainability-enhanced images are fed into a Vision-Language Model (VLM). Chain-of-Thought (CoT) prompting is employed to guide the VLM in generating structured, report-like outputs that mirror clinical documentation. These outputs are designed to help ophthalmologists and eye care professionals by providing interpretable, AI-driven insights that support informed clinical decision making.

The remainder of the paper is organized into five sections. Section 2 provides a comprehensive literature review related to glaucoma detection using explainable deep learning frameworks. Section 3 details the methodology adopted, including preprocessing and the data augmentation, model architectures, model explainability tools, ensemble of models, and clinical simulation using the Google’s Gemini model. Section 4 presents the experimentation results. Section 5 discusses the findings in context of clinical relevance and previous work. Section 6 concludes the paper and suggests possible future research directions.

## 2. Literature Review

Glaucoma is one of the leading causes of irreversible blindness worldwide and has been the subject of extensive research aimed at early detection and intervention. Traditional diagnostic methods such as intraocular pressure (IOP) measurement, visual field testing, and optical coherence tomography (OCT), have proven effective but are often time consuming, resource intensive, and require specialized clinical expertise. In recent years, advances in medical imaging and artificial intelligence (AI), particularly deep learning, have significantly transformed the landscape of glaucoma detection. This section provides a critical review of previous work in the field, focusing on imaging modalities, feature extraction techniques, and the integration of machine learning algorithms for automated glaucoma diagnosis.

### 2.1. Deep Learning with Fundus Images for Glaucoma Classification

There are many studies that had used deep learning models for the classification of glaucoma using fundus images. Shyamalee et al [13] proposed fine-tuned Inception-v3, VGG19, and ResNet50 on two datasets i.e., RIM-ONE and ACRIMA. The study used 5-fold cross-validation with the split ratio of 70 : 15 : 15 for training, testing, and validation sets respectively. On RIM-ONE dataset, Inception-v3 achieved an accuracy of 96.56% with AUC of 0.98, VGG19 achieved an accuracy of 94.95% with AUC of 0.95, and ResNet50 achieved an accuracy of 95.49% with AUC of 0.97. On ACRIMA dataset, Inception-v3 achieved an accuracy of 98.52% with AUC of 0.99, VGG19 achieved an accuracy of 92.64% with AUC of 0.94, and ResNet50 achieved an accuracy of 95.58% with AUC of 0.96. Elan-govan et al. [15] introduces an 18-layer CNN model with four convolutional, two max pooling, and one fully connected layer for glaucoma detection from fundus images. The model was tested on DRISHTI-GS1, ORIGA, RIMONE2, ACRIMA, and LAG [14] databases achieving high accuracies of 96.64% on ACRIMA and an overall range of 78.32% to 96.64% across datasets, demonstrating robustness against Gaussian and salt-and-pepper noise, particularly on the LAG database. Ghorui et al. proposed a new deep leaning approach for glaucoma classification using color fundus images. The proposed ProspectNet outperformed VGG16 (82.02% accuracy) and DenseNet121 (96.25% accuracy), achieved 96.63% accuracy and 0.991 AUC [16]. Mallick et al. [17] used Vision Transformer (ViT) and Swin Transformer (ST) models, which are both transformer-based models for the classification of glaucoma. The models were evaluated on the combined datasets (RIM-ONE, DRISHTI-GS, and REFUGE). ViT have an accuracy of 96.4% and an AUC of 93.8. ST achieved an accuracy of 96.7% with 97.6 AUC score.

### 2.2. Enhancing Interpretability in Deep Learning Models

It is established that deep learning models due to their limited interpretability are often considered black-box models [18]. Hence, it is necessary to ascertain whether the models are predicting based on known markers. A study by [19] proposed a generic framework to classify fundus images as normal, diabetic retinopathy (DR), age-related macular degeneration (AMD), and glaucoma. The authors trained a XGBoost classifier and used SHAP to interpret the prediction of the model by identifying the influential features. Shyamalee et al [20]. proposed a framework to segment optical disc (OD) and optic cup (OC) using attention U-Net with ResNet50 in fundus images. Another model, modified by the authors, Inception V3 was used for the classification of glaucoma. Grad-CAM and Grad-CAM++ were used to identify significant regions in identifying glaucoma. Zhao et al. [21] proposed a Grad-CAM guided contrastive learning approach for the diagnosis of retinal disease using fundus images. They used instance discrimination and KNN-based discrimination to learn class-aware representations from the unlabelled data. Grad-CAM was used to generate lesion-focused heatmaps.

### 2.3. Vision-Language Models (VLMs) for Glaucoma Diagnosis

Recently, the use of VLMs can be seen for the detection of glaucoma by providing fundus images as input and getting the verdict. One study evaluated GPT-4V for the assessment of glaucoma using 300 images from 3 datasets (ORIGA, ACRIMA, and RIM-ONE). GPT-4V achieved detection accuracies of 0.68, 0.70, and 0.81 respectively, slightly below expert graders (0.78, 0.80, 0.88 for grader 1; 0.72, 0.78, 0.87 for grader 2), with about 35% requiring multiple prompts [**?**]. Another study used GPT-4 on the REFUGE dataset for glaucoma classification without pre-training. It achieved a 90% accuracy (95% CI: 87.06%-92.94%) with a specificity of 94.44% (95% CI: 92.08%-96.81%) and a low sensitivity of 50% (95% CI: 34.51%-65.49%). Pre-processing with cropping improved sensitivity to 87.50% but reduced specificity to 56.52%, while adding CLAHE increased sensitivity to 62.50% with a further drop in specificity to 55.43% [23].

The proposed framework not only enhances the diagnostic performance but also address the interpretability issue by combining deep learning models and visualization heatmaps with generative vision language models for clinically meaningful explanations.

## 3. Methodology

In this study, we have presented a comprehensive glaucoma classification framework that utilizes the strengths of both convolutional and transformer-based architectures. Specifically, five deep learning models are employed, three based on Convolutional Neural Networks (CNNs) and two on Vision Transformers (ViTs), to perform a robust classification of glaucoma from ocular images. To ensure model generalizability and improve performance, extensive data preprocessing and augmentation techniques are applied. Each model is trained independently on the preprocessed dataset, and explainability maps are generated using eXplainable AI (XAI) techniques to provide insight into the decision-making process of each model. To further enhance predictive accuracy and reduce model bias, an ensemble strategy was implemented by combining the outputs of all five models. The ensemble output, along with the corresponding XAI visualizations, is then passed into a Vision-Language Model (VLM). Using manually crafted prompts, the VLM generates human-readable textual interpretations that not only describe the classification outcome but also explain the regions of interest highlighted by the XAI methods. An overview of the proposed methodology is illustrated in Figure 1.

**Figure 1:**
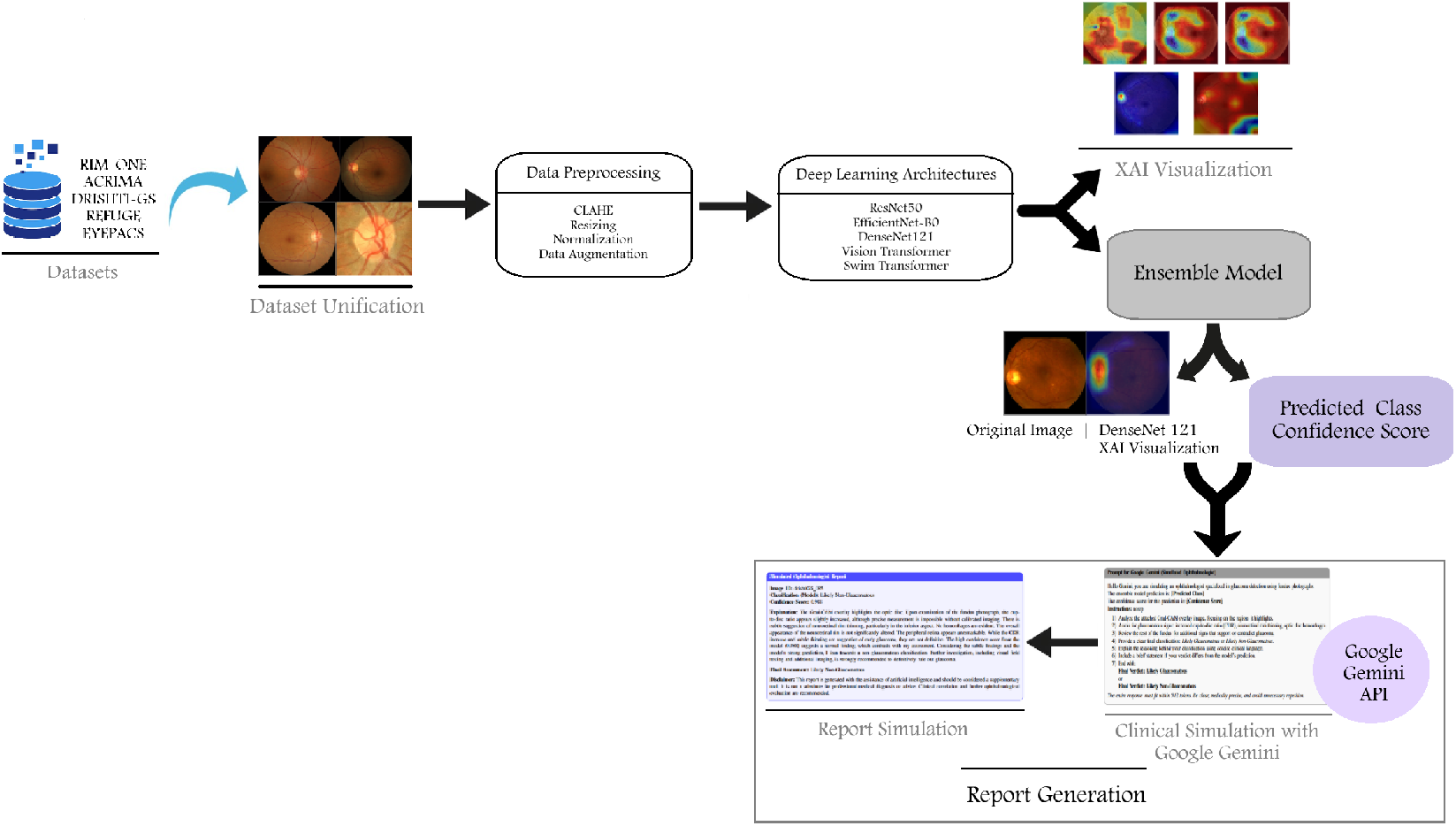
Overview of the glaucoma detection methodology including dataset processing, model training, ensemble inference, and clinical simulation.

### 3.1. Datasets

In this study, five publicly available datasets were used to ensure robust and generalizable glaucoma detection. These datasets provide a diverse pool of labeled images from various sources and populations.

**RIM-ONE DL** dataset contain 485 fundus images having 172 cases of glaucoma and 313 normal cases. It provides ground-truth as well segmentation masks for OD and cup annotated by a specialist for glaucoma assessment [29].

**ACRIMA** dataset contain 705 fundus images with 396 glaucomatous and 309 normal images. Images in ACRIMA database were annotated by two glaucoma experts with 8 years of experience. There are no segmentation masks available for the dataset and can only be used for classification task [25].

**DRISHTI-GS1** dataset contain 151 fundus images. It is organized into 50 training and 51 testing images, with manual segmentations of the optic disk and cup annotated by four experts. It includes structural labels and clinical assessments regarding the presence of glaucoma and notching, thereby facilitating tasks related to OD and cup segmentation and glaucoma diagnosis [26].

**REFUGE** dataset contain 1200 fundus images which are annotated by experts for the segmentation of OD and cup, in addition to clinical labels for glaucoma diagnosis. It serves as a comprehensive benchmark dataset designed to aid in the development and evaluation of algorithms for detecting glaucoma [27].

**EyePACS AIROGS - Light** is a machine-learning-ready glaucoma dataset. The dataset is split into 3 folders: training, validation, and test, which contain 4000, 385, and 385 fundus images in each class respectively. Each training set has a folder for each class: referable glaucoma (RG) and non-referable glaucoma (NRG) [28].

To ensure consistency across diverse datasets and facilitate a robust model training pipeline, a dataset unification script was created. This step aimed to collect all the images from the five datasets and then unify them into one file with standardized file paths and labels. Consistent labelling was enforced by mapping *Glaucoma* to 1 and *Normal* to 0. Only valid image formats (.jpg, .jpeg, and .png) were retained. A metadata CSV file containing images path, labels and dataset names was generated. The final unified dataset was zipped and stored for further use in the model training process. This unification step ensured that all subsequent preprocessing and model training could be applied uniformly across all images, regardless of their original dataset structure or labelling scheme A summary of all the datasets used in this study are provided in Table 1.

**Table 1.** Summary of glaucoma datasets used.

| Dataset | Total Images | Glaucoma | Normal | Reference |
| --- | --- | --- | --- | --- |
| RIM-ONE | 485 | 172 | 313 | [29] |
| ACRIMA | 705 | 396 | 309 | [30] |
| DRISHTI-GS | 50 | 32 | 18 | [26] |
| REFUGE | 800 | 80 | 720 | [31] |
| EYEPACS (Subset) | 9,540 | 4,770 | 4,770 | [32] |
| **Total** | **11,580** | **5,450** | **6,130** | — |

### 3.2. Preprocessing and Data Augmentation

To improve image quality and enhance the generalizability of the models, a series of preprocessing and augmentation techniques were applied. These steps were crucial for ensuring that the models could effectively learn relevant features across diverse patient samples and imaging conditions. One key preprocessing method employed was Contrast Limited Adaptive Histogram Equalization (CLAHE), a sophisticated contrast enhancement technique widely used in medical imaging. Unlike global histogram equalization, CLAHE operates on small regions within the image, enhancing local contrast while preventing over-amplification of noise. In the context of this study, CLAHE was particularly valuable for accentuating subtle variations in the optic disc and cup regions, which are critical for glaucoma detection. By making pathological cues more visually prominent, this enhancement helped facilitate more accurate feature learning and, ultimately, more reliable model predictions.

To ensure consistency across model architectures and enable efficient training, all input images were resized according to the requirements of the respective networks: CNN-based models received inputs standardized to 512 × 512 pixels, while transformer-based models were resized to 224 × 224 pixels, in line with common Vision Transformer input dimensions. Normalization was performed using the standard ImageNet mean and standard deviation values, a widely adopted practice that facilitates effective transfer learning and promotes training stability by aligning input distributions with pre-trained model expectations.

During training, data augmentation was employed exclusively to improve model robustness and reduce overfitting. The augmentation pipeline included random horizontal flipping, brightness and contrast adjustments, and random rotations, which introduced controlled variability in the training data while preserving the clinical relevance of the images. These transformations were implemented using the Albumentations library and seamlessly integrated into the workflow through a custom PyTorch Dataset class, ensuring consistent and reproducible application of augmentations during each training epoch.

### 3.3. Deep Learning Ensemble Modeling

#### 3.3.1. CNN-Based Models

Convolutional Neural Networks (CNNs) have demonstrated remarkable success in medical image classification tasks, particularly due to their ability to capture and learn hierarchical spatial features. In this study, three well-established CNN architectures ResNet50, EfficientNet-B0, and DenseNet121 were employed for automated glaucoma detection. These models were chosen based on their proven efficacy in medical imaging applications, offering a balance between depth, parameter efficiency, and representational power.

ResNet50, a 50-layer deep residual network with approximately 23 million trainable parameters [33], provides a lightweight yet powerful alternative by employing a compound scaling method that uniformly scales depth, width, and resolution. EfficientNet-B0 is a fundamental model from the family of EfficientNet. It has 5.29 million trainable parameters [34]. DenseNet121, comprising 7 million parameters [35], encourages feature reuse by densely connecting layers, which is particularly beneficial for detecting subtle pathological cues in optic nerve head regions.

All CNN models were initialized with ImageNet pretrained weights to use strength of transfer learning, which accelerated convergence and enhanced model generalization to the glaucoma classification task. Preprocessed fundus images enhanced using CLAHE for improved contrast in the optic disc and cup areas and normalized with ImageNet statistics—were fed into these models. Each model performed binary classification, predicting the presence or absence of glaucoma.

Training was conducted using the cross-entropy loss function, optimized via the AdamW optimizer with an adaptive learning rate scheduler to balance learning speed and generalization. To evaluate model performance comprehensively, key metrics such as accuracy, precision, recall, F1-score, and ROC-AUC were monitored across training and validation sets. Additionally, to ensure transparency and interpretability in clinical decision-making, Grad-CAM was employed to visualize salient regions in the input images that significantly influenced each model’s predictions, offering visual insights into the learned discriminative patterns.

#### 3.3.2. Transformer-Based Models

While originally developed for natural language processing tasks, transformer architectures have recently achieved state-of-the-art performance in various vision applications, owing to their ability to model long-range dependencies and contextual relationships more effectively than traditional convolutional networks. In this study, we explored two transformer-based models for the task of glaucoma detection: the Vision Transformer (ViT-Base Patch16 224) and the Swin Transformer (Swin-Base Patch4 Window7 224). These architectures were selected due to their distinct design paradigms that offer complementary strengths in visual representation learning.

The ViT-Base Patch16 224 model operates by dividing input images into non-overlapping patches of size 16 × 16, which are then linearly embedded and treated as to-kens—analogous to words in NLP. These patch embeddings are passed through a series of self-attention layers that learn global relationships across the entire image [36]. This global context modeling is particularly useful for detecting glaucoma-related changes that may not be localized to a specific region but instead involve subtle variations across the optic disc, cup, and surrounding retinal structures.

On the other hand, the Swin Transformer (Shifted Windows Transformer) introduces a more localized and hierarchical approach to visual processing. It begins by splitting the image into smaller patches (via Patch4) and applies self-attention within local windows of size 7, which are gradually shifted between layers to enable cross-window information flow [37]. This design allows the model to efficiently capture both fine-grained local details and broader contextual information, which is critical in medical imaging scenarios where multi-scale feature extraction can greatly influence diagnostic accuracy.

Both transformer models were pretrained on ImageNet and subsequently fine-tuned on the glaucoma classification dataset, with their classification heads adapted for binary classification (glaucoma vs. non-glaucoma). The training protocols—including optimizer, learning rate scheduling, and loss function—were aligned with those used for the CNN models to ensure comparability in performance.

To enhance interpretability and gain insight into model decision-making, we employed attention rollout visualization techniques. These methods aggregate attention weights across all transformer layers and heads to produce heatmaps that reveal the regions of the image that most influenced the model’s predictions. This transparency is especially important in medical AI applications, where explainability can support clinical validation and trust in automated diagnosis.

### 3.4. Performance Metrics

To rigorously assess the effectiveness of the proposed models in glaucoma detection, we employed a set of widely accepted performance metrics: accuracy, precision, recall, F1-score, and area under the curve (AUC). These metrics not only measure the overall correctness of the models but also provide deeper insight into their sensitivity to positive cases and resistance to false alarms—critical aspects in the context of medical diagnosis.

Accuracy indicates the proportion of total correct predictions, offering a general sense of how well the model performs across all classes. However, accuracy alone may not be sufficient in medical imaging tasks where class imbalance is common (e.g., fewer glaucoma-positive cases compared to healthy ones). It is defined as:

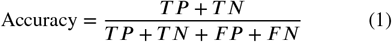

Precision measures the ability of the model to correctly identify only relevant instances. In the glaucoma detection context, it reflects the proportion of cases predicted as glaucoma that are actually glaucoma, helping to reduce false positives.

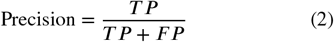

Recall (or sensitivity) evaluates how effectively the model captures true positive cases. A high recall is essential to avoid missing glaucoma cases, as early diagnosis is vital for preventing vision loss.

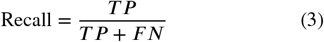

F1-score is the harmonic mean of precision and recall, providing a balanced measure when both false positives and false negatives are of concern an ideal metric for healthcare settings where both types of errors can be costly.

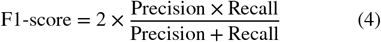

AUC (Area Under the Receiver Operating Characteristic Curve) offers a threshold-independent measure of model performance by quantifying its ability to distinguish between glaucomatous and non-glaucomatous cases. A higher AUC indicates better discrimination capability, even across varying decision thresholds.

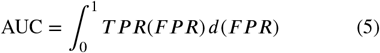

By using this comprehensive set of metrics, we ensured a robust evaluation of the models’ performance not just in terms of overall accuracy, but also their clinical reliability in correctly identifying glaucoma patients with minimal diagnostic error.

## 4. Interpretable Analysis of Glaucoma Images

To ensure transparency and gain deeper insight into the decision-making processes of the deep learning models used for glaucoma detection, two state-of-the-art explainability techniques were employed, each specifically tailored to the underlying architecture of the models. In clinical applications, interpretability is essential not only for validating the correctness of predictions but also for building trust among healthcare professionals.

*Gradient-weighted Class Activation Mapping (Grad-CAM)* was utilized for all CNN-based models (ResNet50, EfficientNet-B0, and DenseNet121). Grad-CAM [38] generates heatmaps by using the gradients of the predicted class flowing into the final convolutional layer to identify spatial locations in the input image that strongly influence the prediction. This allows for visual interpretation of the model’s focus—highlighting anatomical regions such as the optic disc and cup that are clinically relevant in glaucoma diagnosis. These visual cues provide critical feedback on whether the model is learning medically meaningful features or relying on spurious patterns.

*Attention Rollout* was applied to transformer-based models (ViT-Base and Swin Transformer), which rely on self-attention mechanisms instead of convolution. Attention Rollout [39] aggregates attention weights across all transformer layers and heads, tracing how information flows from the input patches to the final decision. This results in an attention map that reveals which regions of the image the model “attended to” most during inference. A final attention mask was derived by computing the influence of the input patches on the classification token, which was then reshaped into a 2D grid that matched the input structure. The mask was normalized and overlaid as a heatmap on the corresponding fundus image. Given that transformers lack an inherent spatial bias, this visualization is particularly important to confirm whether the model is attending to diagnostically important structures in the image.

Together, these techniques offer complementary insights into model interpretability Grad-CAM for convolution-based spatial learning and Attention Rollout for attention-driven contextual modeling. Their integration improves the credibility of automated glaucoma detection systems by aligning machine decisions with known medical reasoning, thus supporting more informed clinical adoption.

### 4.1. Ensemble Inference

To ensure a robust and reliable classification of glaucoma, all five trained models were utilized during the inference phase. Each model was loaded with its corresponding fine-tuned weights and executed in evaluation mode to maintain consistent behavior and prevent any updates to internal parameters such as batch normalization statistics. The input resolution was dynamically selected based on the model architecture: CNN-based models processed images resized to 512×512, while transformer-based models received input of 224 × 224, according to their design specifications.

During inference, each model produced softmax normalized class probabilities, representing the likelihood of the input image belonging to either the glaucomatous or non-glaucomatous class. To derive a consensus prediction and take advantage of the complementary strengths of all models, these probability distributions were averaged across all five models. The class with the highest average probability was selected as the final prediction, and this value was also recorded as the confidence score of the model. This ensemble-based decision strategy not only improved the reliability of the prediction, but also reduced the variance associated with the output of any single model.

A Grad-CAM heatmap was generated using one of the top-performing CNN-based models to visualize the regions of the input image that most influenced the model’s prediction. To facilitate interpretability in a clinical context, the heatmap was concatenated side-by-side with the original image, producing a combined 1024×512 visualization. The composite image was then utilized in the subsequent clinical simulation step to support visual analysis and decision-making by healthcare professionals.

### 4.2. Clinical Simulation via Gemini API

To emulate expert-level clinical interpretation, we integrated a vision-language reasoning step using Google’s Gemini 1.5 Flash model, accessed via the google-generativeai Python SDK. This step was designed to bridge the gap between the raw model outputs and human-readable clinical insights. A carefully structured prompt was created, which incorporated the predicted label of the ensemble model, the corresponding confidence score, and a composite image that combined the original fundus photograph with its Grad-CAM heatmap visualization.

The prompt explicitly instructed Gemini to evaluate hallmark indicators of glaucoma—such as optic disc cupping, neuroretinal rim thinning, and the presence of optic disc hemorrhages—based on visual cues in the input. The model was then asked to provide a concise clinical rationale followed by a final assessment in one of two categories: Likely Glaucomatous or Likely Non-Glaucomatous. To maintain clinical relevance and avoid unnecessary verbosity, the response was constrained to 512 tokens, ensuring clarity and interpretability for real-world diagnostic scenarios. The complete prompt template used in this simulation is illustrated in Figure 2.

**Figure 2:**
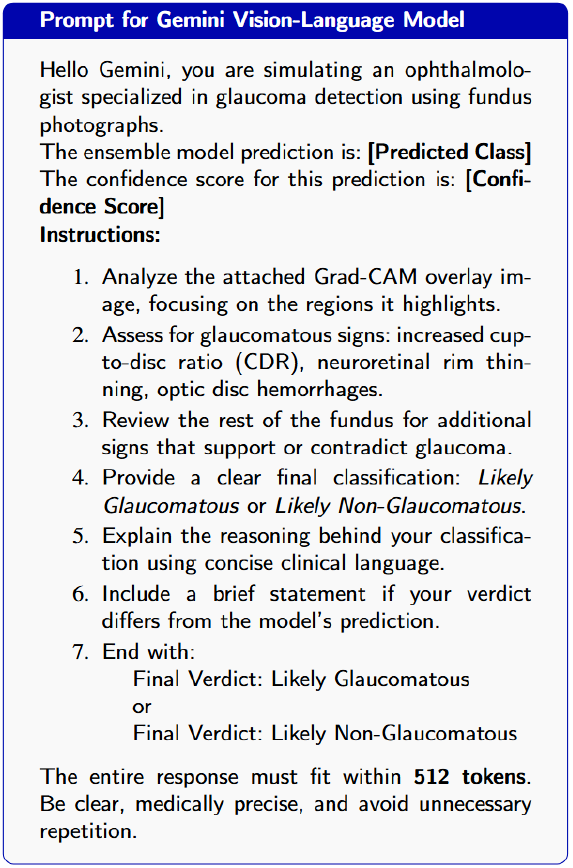
Prompt provided to Gemini for simulating an ophthalmologist’s clinical reasoning in glaucoma detection.

## 5. Experimental Analysis

To assess the effectiveness of deep learning approaches for the detection of glaucoma from fundus images, a total of five pretrained models comprising three CNN-based architectures and two transformer-based architectures were fine-tuned using transfer learning on a labeled dataset. Using pretrained weights allowed the models to adapt more efficiently to the medical imaging domain while reducing training time and data dependency. Each model was trained for up to 25 epochs, with early stopping implemented based on the validation F1-score to prevent overfitting and ensure optimal generalization. Model performance was comprehensively evaluated using key classification metrics, including accuracy, precision, recall, F1-score, loss, and Area Under the ROC Curve (AUC), providing a balanced and clinically relevant assessment of each model’s diagnostic capability.

### 5.1. Models Evaluation

#### ResNet50 Performance

The ResNet50 model demonstrated consistent performance improvements over epochs, achieving its based validation F1-score of 0.9426 at epoch 19. The final model reported the metrics in Table 2.

**Table 2.** ResNet50 Model Performance Metrics Across Training and Validation Datasets.

| Metrics | Train | Validation |
| --- | --- | --- |
| Loss | 0.0353 | 0.1962 |
| Accuracy | 0.9868 | 0.9434 |
| Precision | 0.9869 | 0.9379 |
| Recall | 0.9851 | 0.9422 |
| F1-score | 0.9860 | 0.9400 |
| AUC | 0.9992 | 0.9843 |

The visualization of all performance metrics across the dataset is provided in Figure 3a.

**Figure 3:**
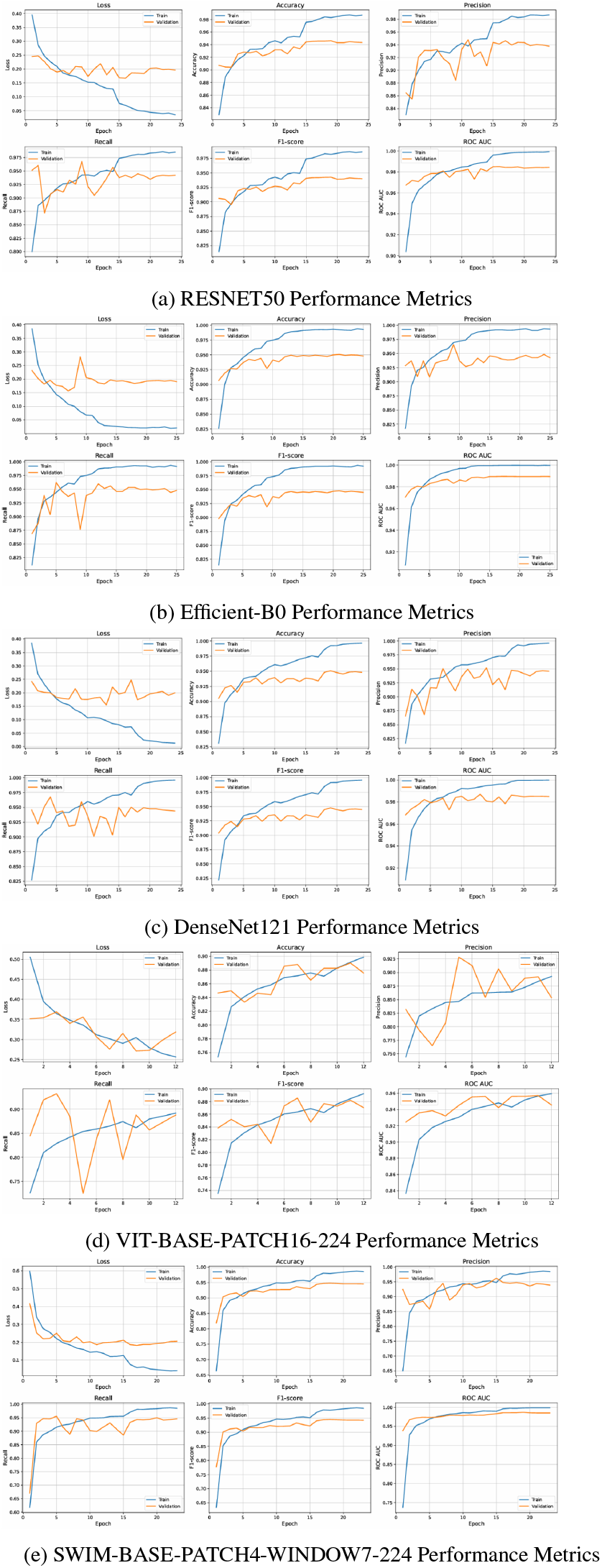
Visualization of training and validation performance metrics (Loss, Accuracy, Precision, Recall, F1-score, and AUC)

ResNet50 demonstrated strong performance in the classification of glaucoma with a validation accuracy of 94.56% and an AUC of 0.9849, indicating high discriminative capability. The model achieved an F1-score of 0.9424, showing a good balance between precision (93.90%) and recall (94.59%). These results suggest that ResNet50 is effective in correctly identifying images of glaucomatous and normal fundus images with minimal misclassification. The corresponding confusion matrix (Figure 7a) shows the normalized prediction distribution, where values represent proportions rather than raw counts.

#### EfficientNet-B0 Performance

EfficientNet-B0 achieved the best validation F1-score of 0.9478 at epoch 21. This model showed excellent generalization with a good balance between training and validation performance. The final accuracies of the model are reported in Table 3. The visualization of all performance metrics across the dataset is provided in Figure 3b.

**Table 3.** EfficientNet-B0 Model Performance Metrics Across Training and Validation Datasets.

| Metrics | Train | Validation |
| --- | --- | --- |
| Loss | 0.0201 | 0.1906 |
| Accuracy | 0.9928 | 0.9482 |
| Precision | 0.9931 | 0.9425 |
| Recall | 0.9915 | 0.9477 |
| F1-score | 0.9923 | 0.9451 |
| AUC | 0.9997 | 0.9894 |

EfficientNet-B0 achieved the highest performance among the CNN-based models, with a validation accuracy of 94.96%, F1-score of 0.9448, and an AUC of 0.9849. Its precision (94.74%) and recall (94.31%) were well-balanced, indicating that the model maintained both sensitivity and specificity. The model’s lightweight architecture combined with strong performance makes it particularly suitable for deployment in resource-constrained settings. Figure 7b illustrates its classification results.

#### DenseNet121 Performance

DenseNet121 reached a best validation F1-score of 0.9449 and AUC of 0.9999. However, it achieved this with a slightly better training metrics and a lower training loss, indicating highly efficient learning without overfitting. Table 4 shows the metrics for DenseNet121.

**Table 4.** DenseNet121 Model Performance Metrics Across Training and Validation Datasets.

| Metric | Train | Validation |
| --- | --- | --- |
| Loss | 0.0128 | 0.1998 |
| Accuracy | 0.9964 | 0.9482 |
| Precision | 0.9961 | 0.9458 |
| Recall | 0.9963 | 0.9440 |
| F1-score | 0.9962 | 0.9449 |
| AUC | 0.9999 | 0.9850 |

The visualization of all the performance metrics across the dataset are provided in Figure 3c. DenseNet121 also performed robustly, achieving a validation accuracy of 94.35%, an F1-score of 0.9405, and an AUC of 0.9840. The model maintained a recall of 94.59%, suggesting a strong ability to correctly detect glaucoma cases, while its precision of 93.55% ensured minimal false positives. These metrics reflect DenseNet121’s ability to generalize well, even under subtle variations in the fundus images. The confusion matrix in Figure 7c presents the model’s prediction outcomes and shows the normalized prediction distribution, where values represent proportions rather than raw counts.

The two pretrained transformer-based models used in this study were ViT Base Patch16 224 and Swin Base Patch4 Window7 224. The performance of all these models is provided below.

#### Vision Transformer (ViT Base Patch16 224) Performance

The Vision Transformer model was trained using 25 epochs with early stopping enabled. It achieved a validation F1-score of 0.8705, with corresponding accuracy of 87.56%, precision of 85.36%, and recall of 88.81%. The model demonstrated an AUC of 0.9457, indicating strong discriminative ability in glaucoma detection from fundus images. The training performance was also strong, with an F1-score of 0.8924 and AUC of 0.9596. Despite its transformer-based architecture, which is less inductive-biased than CNNs, ViT performed comparably well. However, it showed slightly lower generalization than CNN-based models. The training converged by epoch 12 due to no further improvement in validation F1-score. The visualization of all the performance metrics across the dataset are provided in Figure 3d. The model demonstrated slightly more false positives (glaucoma predicted as normal) than false negatives, consistent with its higher recall. This balance suggests a cautious classification approach, which favors sensitivity to the presence of disease. The normalized confusion matrix is given in Figure 7d.

#### Swin Transformer (Swin Base Patch4 Window7 224) Performance

Swin Transformer outperformed all other models, including CNN-based and standard transformer models, achieving the highest validation F1-score of 0.9448. It also attained a validation accuracy of 94.56%, precision of 93.90%, recall of 94.59%, and an AUC of 0.9849. The model benefited from hierarchical feature representation and local window-based attention mechanisms, which likely contributed to its superior performance. On the training set, Swin Transformer recorded an F1-score of 0.9848 and an exceptional AUC of 0.9985, suggesting minimal overfitting and high confidence in predictions. The training progressed for 23 epochs before early stopping was triggered due to stagnation in validation F1.

The visualization of all the performance metrics across the dataset are provided in Figure 3e. The normalized confusion matrix revealed balanced performance with low mis-classification rates for both glaucoma and normal classes. All training and validation metrics (loss, accuracy, precision, recall, F1-score, and AUC) were plotted over epochs to visualize the model’s learning behaviour and stability. The normalized confusion matrix is shown in Figure 7e.

### 5.2. Visual Interpretation

To understand the model decisions, visual interpretation techniques were applied to both CNN-based and transformer-based models. Grad-CAM was used for CNN-based models, while attention rollout was applied to transformer-based models. A total of ten fundus images were analysed, five from glaucoma cases and five from normal cases.

**ResNet50**, in glaucoma images, showed dispersed attention and failed to accurately localize the disc and cup in most cases (Figure 5a). In normal images, attention appeared consistently in a region near the disc but did not align precisely with it, possibly focusing on areas with patterns commonly seen in normal cases.

**Figure 4:**
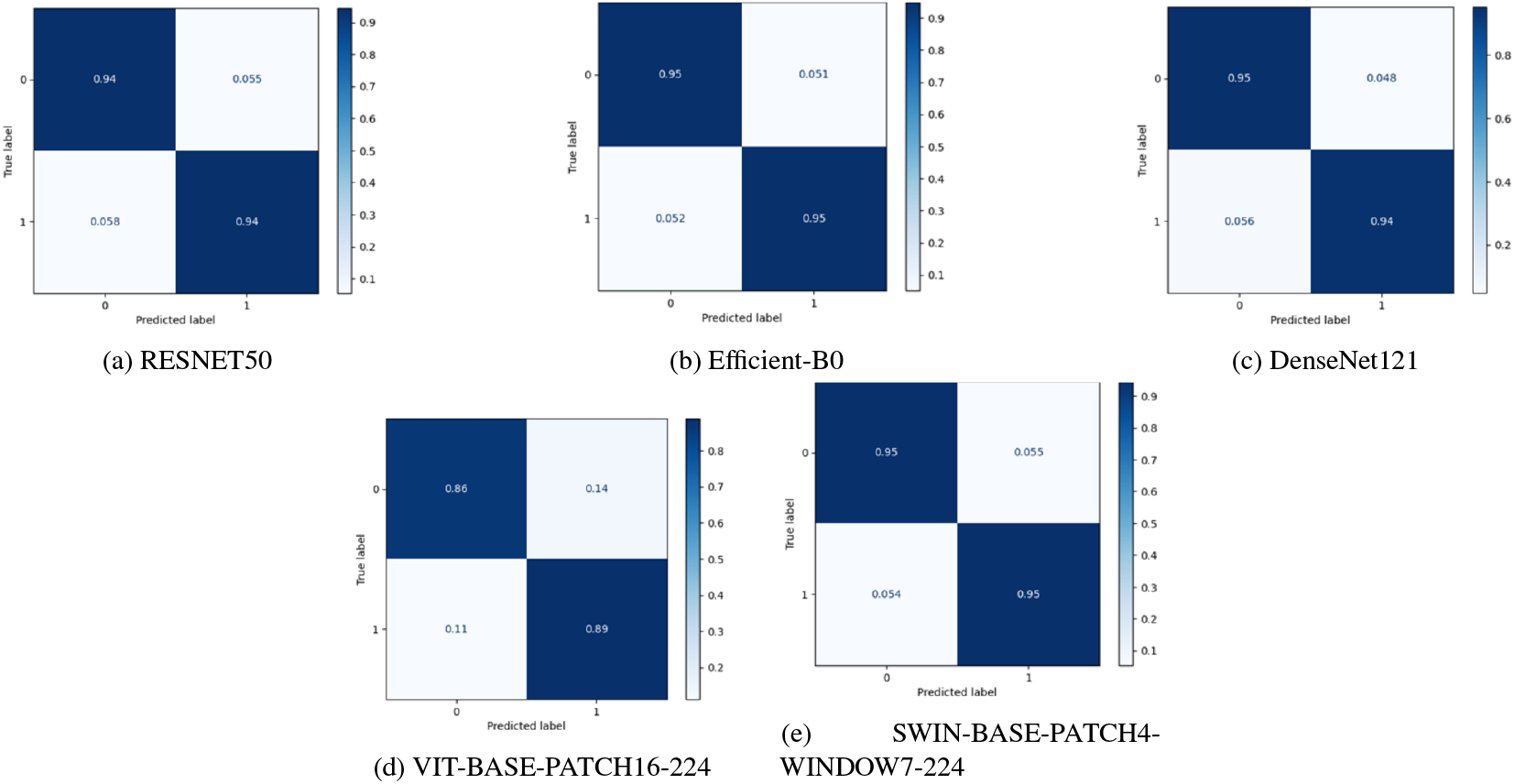
Normalized Confusion Matrices for Various Models

**Figure 5:**
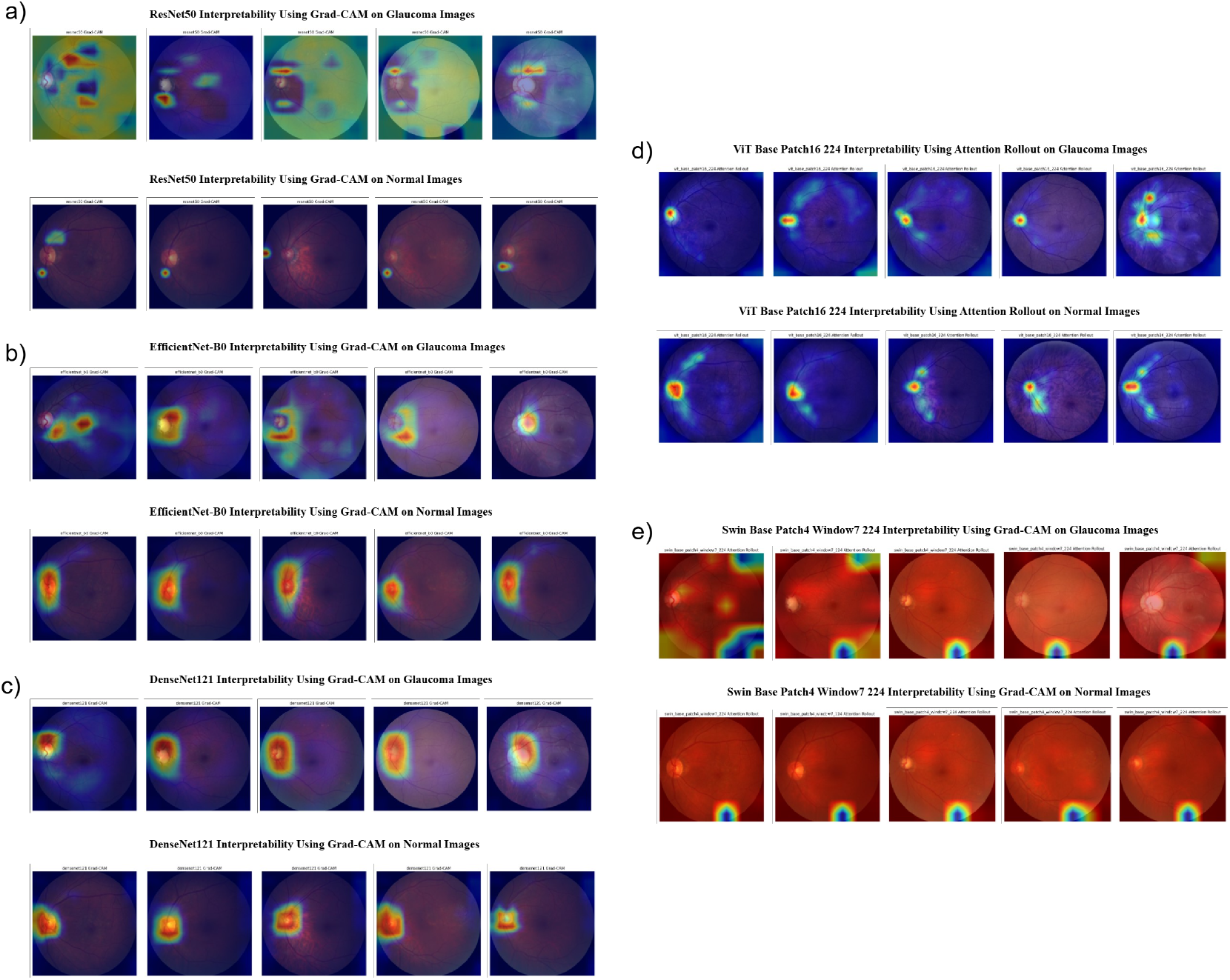
Grad-CAM results on CNN-based model predictions and attention rollout visualizations on transformer-based predictions. a) Grad-CAM results on ResNet50 model predictions. b) Grad-CAM results on EfficientNet-B0 model predictions. c) Grad-CAM results on DenseNet121 model predictions. d) Grad-CAM results on ViT model predictions. e) Grad-CAM results on Swin transformer model predictions.

**EfficientNet-B0** model, in glaucoma images, mostly failed to highlight the disc and cup accurately (Figure 5b), with attention appearing only near or partially over the disc in a few cases. In normal images, the attention was consistently centered on the disc region. This may suggest the model is focusing on familiar patterns linked to normal anatomy.

**DenseNet121** model, in both glaucoma and normal images, consistently focused attention on the disc region (Figure 5c). Most images showed red and yellow activation directly over the disc, with a few highlighting the disc border. This indicates strong and consistent localization of relevant anatomical features.

**ViT Base Patch16 224** showed consistent concentric attention patterns across all glaucoma and normal images, focusing on the disc region with red at the center, surrounded by yellow and green. The rest of the image appeared mostly blue, indicating low attention outside the disc. This suggests the model is specifically attending to the disc region, which is a critical anatomical feature common to both classes. The focused red attention on the disc implies that ViT is localizing important regions rather than diffusing attention across the entire image. The visualization results are shown in Figure 5d.

**Swin Base Patch4 Window7 224** showed red attention across the entire image in both glaucoma and normal cases, with layered blue, green, and yellow outlines appearing randomly near the bottom or edges. These patterns did not correspond to the disc or cup region and lacked consistency across images. This suggests the model may not have learned to focus on relevant anatomical features and might be relying on unrelated visual cues. The results are shown in Figure 5e. Ensemble model which was constructed by averaging the probabilities of all the five models. The ensemble model was evaluated on a test dataset showed AUC of 0.99. The AUC-ROC graph is present in the Figure 6a. The confusion matrix in Figure 6b illustrates the performance of the ensemble model on the test dataset. Out of the total normal samples, 1170 were correctly classified as normal, while 56 were mis-classified as glaucoma. Similarly, out of the glaucoma cases, 1039 were correctly identified, with only 51 misclassified as normal. This strong diagonal dominance in the matrix indicates high classification accuracy and robust discriminative capability of the model in distinguishing between normal and glaucomatous images. The performance of ensemble model on test data is shown in Table 7.

**Table 5.** ViT Base Patch16 224 Model Performance Metrics Across Training and Validation Datasets.

| Metrics | Train | Validation |
| --- | --- | --- |
| Loss | 0.2567 | 0.3190 |
| Accuracy | 0.8987 | 0.8756 |
| Precision | 0.8928 | 0.8536 |
| Recall | 0.8920 | 0.8881 |
| F1-score | 0.8924 | 0.8705 |
| AUC | 0.9596 | 0.9457 |

**Table 6.** Swin Base Patch4 Window7 224 Model Performance Metrics Across Training and Validation Datasets.

| Metrics | Train | Validation |
| --- | --- | --- |
| Loss | 0.0410 | 0.2057 |
| Accuracy | 0.9856 | 0.9456 |
| Precision | 0.9842 | 0.9390 |
| Recall | 0.9853 | 0.9459 |
| F1-score | 0.9848 | 0.9424 |
| AUC | 0.9985 | 0.9849 |

**Table 7.** Performance metrics for the proposed ensemble model.

| Metrics | Score |
| --- | --- |
| Accuracy | 0.9538 |
| Precision | 0.9489 |
| Recall | 0.9532 |
| F1-score | 0.9510 |
| AUC | 0.9909 |

**Figure 6:**
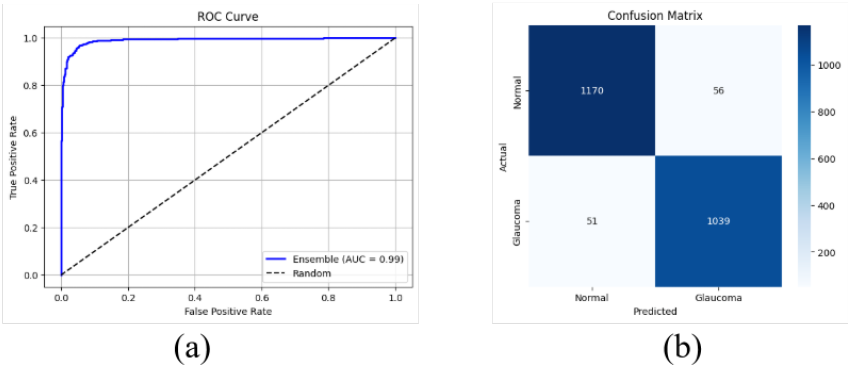
ROC curve of the ensemble model on the test dataset.

**Figure 7:**
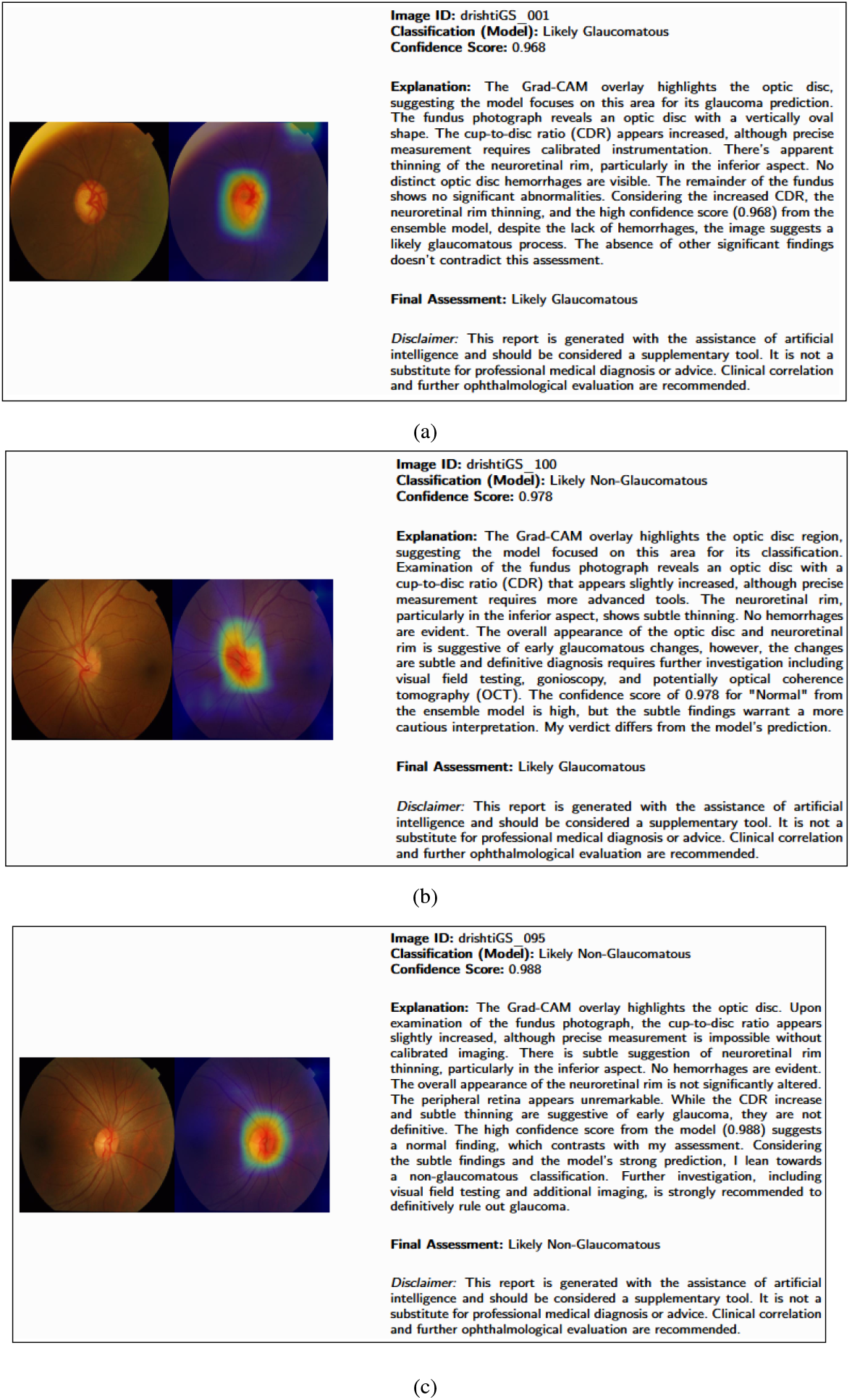
Simulated Ophthalmologist Report

After the ensemble model has given its prediction, a Grad-CAM image from the DenseNet121 (best Grad-CAM outputs) is merged with the original fundus image. The image with the outcomes of the ensemble model is provided to the Google’s Gemini 1.5 flash which is prompted in a fashion that it takes the image id, image, predicted class. It outputs the explanation by first analyzing the Grad-CAM output and then it analyzes the original fundus image in order to look for other signs involved in the glaucoma. At the end it gives a final verdict based on the explanation. The output for a glaucoma diagnosed image is provided in Figure **??**a. There are some times when the test data label and the Gemini model’s inference has a difference. In that case Gemini’s final verdict is different then the ensemble prediction. An example of thisis shown in Figure **??**b. The output for a normal case is shown in Figure **??**c.

## 6. Results and Discussion

This study demonstrated the use of ensemble deep learning models using 5 deep learning architectures, 3 CNN-based and 2 transformer-based. All 5 models were trained on the publicly available datasets and validated. Ensmeble model was built using the deep learning models and was validated on the test dataset. The ensemble model gave the accuracy of 86.27% with an AUC of 0.8927. Grad-CAM image from the DenseNet121 was generated because it always generated the heatmaps highlighting the most significant parts when classifying glaucoma, optic disc and optic cup. There are a few previous studies that have employed the use of enesmble models. However, most of them have used the ensemble model for segmentation tasks. One study has used ensemble of 3 CNN models to classify glaucoma using REFUGE dataset. Individual models showed the maximum AUC of 0.92 and maximum accuracy of 97%. The ensemble methods used in the study include hard-voting and average voting. Average voting ensemble method gave the maximum accuracy among all the models of 98% with an AUC of 0.88 [40].

One of the works by [41] used datasets that is not available publicly for glaucoma classification. The study used ensemble of 56 CNN models and 4 CNN architectures (including InceptionNet-V3 and InceptionResNet-V2 with varied fully connected layers). Their approach acheived an average AUROC of 0.975 and accuracy of 0.881 compared to 0.950 AUROC and 0.852 accuracy from the individual best-performing CNN model. Our ensemble model on the validation data gave the accuracy of 86.27% with an UC of 0.8927. It is because we have done testing on the test dataset of DRISHTI-GS dataset which has only 51 fundus images while it is trained on 50 of its train images. Our models were trained on 5 different datasets having different types of fundus images. Moreover, our work entailed the use of a vision language model, Google Gemini 1.5 flash, to generate a report having an explanation of the predicted class and the Grad-CAM result produced. Google Gemini 1.5 flash can take audio, video, images, and text as inputs and outputs text. The Gemini model, accessed via the Google Gemini API, was selected for its versatility and strong performance across a wide range of vision-language tasks. Its integration into our pipeline was streamlined through a modular setup—allowing the model to be easily swapped by simply modifying the model name in the API call. This flexibility ensures future-proofing of the system, enabling seamless updates in the event of model deprecation or the availability of improved alternatives. In our workflow, Gemini was provided with a structured prompt containing the ensemble model’s predicted label, the associated confidence score, and a composite image combining the original fundus image with its corresponding Grad-CAM heatmap. The prompt instructed the model to interpret this information and generate a clinically meaningful report-style output, designed specifically for clinicians who may not have expertise in reading heatmaps. This approach bridges the gap between complex model outputs and human-understandable explanations, supporting more accessible and trustworthy clinical decision-making.

While recent studies have begun exploring the use of large language models such as OpenAI’s ChatGPT, Anthropic’s Claude, and BERT—for medical diagnosis, these models are often applied in a more generalized manner, attempting to infer glaucoma directly from textual input or limited visual cues [42]. In contrast, our method takes a more informed and structured approach, by providing both the model’s predicted outcome and its explanation (via Grad-CAM) as input to the language model. This enhances the interpretability and reliability of the generated output, aligning the process more closely with clinical reasoning.

### 6.1. Comparative Analysis

Most recent works include the binary classification to differentiate between glaucoma-affected eyes and normal eyes. However, these studies do not have a large amount of data available. Moreover, not every work includes the explainability of the models using explainable AI tools such as Grad-CAM, etc. In our work, we have not only trained deep learning models but we have also constructed an ensemble model to enhance the prediction robustness. Furthermore, we introduced a novel integration of Google Gemini 1.5 flash vision-language model to generate clinically coherent textual explanations.

Anwar et al. proposed an ensemble model for binary classification of normal and advanced glaucoma using the Harvard dataverse dataset. Their model acheived an overall accuracy of 98.04% [43]. Other notable works that have used ensemble models employing deep learning are presented in the Table 8.

**Table 8.** Comparison of this study with other notable works.

| Reference | Dataset | Validation Accuracy (%) | Sensitivity (%) | Specificity (%) |
| --- | --- | --- | --- | --- |
| [44] | DRIONS-DB, HRF, DRISHTI-GS, PSGIMSR, RIM-One | 88.96 | 81.00 | 95.53 |
| [45] | REFUGE, ACRIMA | 95.30 | 95.00 | 99.00 |
| [46] | Drishti-GS, RIM-One | 97.51 | 98.78 | 96.20 |
| [47] | Harvard Dataverse, RIM-One | 94.45 | 95.20 | 93.72 |
| [43] | Harvard Dataverse | 98.04 | 97.00 | 100.00 |
| **Proposed Methodology** | **RIM-ONE, ACRIMA, DRISHTI-GS, REFUGE, EYEPACS** | **95.38** | **95.32** | **95.42** |

### 6.2. Limitations

While the proposed ensemble model has high accuracy and AUC values, there are some limitations in this study. The study used 5 different datasets. All datasets had different numbers of samples in them. This imbalance may have introduced bias during model training and evaluation.

Although the ensemble approach improves robustness by combining multiple models, it also increases computational complexity, which may limit its applicability in real-time or resource-constrained environments. External validation on more diverse and larger cohorts is necessary to further confirm the reliability and clinical utility of the proposed ensemble model. The Grad-CAM visualizations provide relatively coarse heatmaps which may not fully capture subtle or small lesions critical for diagnosis. The quality and consistency of fundus images across different datasets can affect both model performance and the clarity of visual explanations. Furthermore, the report generation via Google Gemini relies on the quality of input explanations and may sometimes produce generic or non-specific outputs without detailed clinical context. Lastly, the integration has not yet been tested extensively in clinical workflows, limiting understanding of its real-world usability and impact.

### 6.3 Future Work

Future research could focus on enhancing the interpretability and clinical applicability of the ensemble model by integrating more advanced explainability techniques beyond Grad-CAM, such as guided Grad-CAM or integrated gradients, to achieve finer localization of pathological features in fundus images. Additionally, expanding the dataset with more diverse populations and imaging devices would improve model robustness and generalizability. The current use of Google Gemini for automated report generation is promising; future work could explore deeper integration with electronic health record systems to provide real-time, patient-specific diagnostic summaries and recommendations. Moreover, investigating how the ensemble model’s predictions and explainability outputs influence clinical decision-making and patient outcomes would be valuable to establish practical utility.

## 7. Conclusion

This study presents a comprehensive and explainable framework for glaucoma detection using fundus images, addressing key challenges in automated diagnosis. By unifying five diverse datasets, we enhanced the generalizability of our approach across varied populations and imaging conditions. The ensemble of five deep learning models (ResNet50, EfficientNet-B0, DenseNet121, Vision Transformer, and Swin Transformer) demonstrated superior performance, achieving a test accuracy of 95.38% and an AUC of 0.99, surpassing individual model results and previous studies. The integration of Grad-CAM and attention rollout visualizations provided transparent insights into model decision-making, consistently highlighting diagnostically relevant regions such as the optic disc and cup. Furthermore, the novel use of Google’s Gemini 1.5 Flash to generate clinician-style diagnostic reports bridged the gap between complex AI outputs and human-readable interpretations, enhancing clinical relevance. Despite these advancements, limitations such as dataset imbalance and coarse heatmap granularity suggest areas for improvement. Future work should focus on incorporating advanced explainability techniques, expanding dataset diversity, and validating the framework in clinical settings to ensure real-world applicability. This approach not only improves diagnostic accuracy but also fosters trust in AI-driven glaucoma detection, paving the way for its integration into ophthalmic practice.

## Data Availability

All data produced are available online at https://github.com/Document-Data-Analyst/Glaucoma-Disease-Detection

https://github.com/Document-Data-Analyst/Glaucoma-Disease-Detection

## Acknowledgment

This research was supported by competitive financial support from the MITACS Globalink Research Internship, Canada, awarded to Syed Ali Raza Naqvi.

